# Model Development and Real-World Deployment of Multimodal Input-Based Subtyping of Depression in Tele-Counseling for Scalable Mental Health Assessment

**DOI:** 10.64898/2026.02.11.25342657

**Authors:** Amal Jude Ashwin Francis, Ahmad Raza, Nischay Patel, Rubal Gajbhiye, Vaneesha S Kumar, Aneesh T, Amandip Saikia, Oyin Mibang, Vidhyadharan K, Kritika Joshi, Lazar Tony, Pragathi Priyadharsini Balasubramani

**Author notes:** equal second authors (Ahmad Raza, Nischay Patel, Rubal Gajbhiye, Vaneesha S Kumar). equal third authors (Aneesh T, Amandip Saikia, Oyin Mibang, Vidhyadharan K, Kritika Joshi, Lazar Tony).

## Abstract

The rapid growth of tele-counseling and the use of lay counselors in high-volume, low-resource mental health services has created a need for scalable tools for early detection and triage. Effective personalization now requires stratifying individuals by dominant symptom profiles, such as appetite, agency, anxiety, and sleep disturbances. Depression symptoms vary widely, even among those with similar scores, reflecting distinct psychophysiological and cognitive-affective patterns. In tele-mental-health settings, where contextual cues are limited, multimodal behavioral signals from natural interactions can complement traditional assessments. Using synchronized audio, video, and text data from the EDAIC dataset (N=275), we propose a multimodal learning framework to classify five clinically validated outcomes: Depression, Appetite disturbance, Agency impairment, Anxiety, and Sleep problems.

We developed a comprehensive multimodal machine-learning pipeline, incorporating automated dataset construction, modality-specific feature extraction (acoustic, facial action unit, linguistic), and supervised learning with cross-validation. Labels were derived from validated scoring rules to ensure clinical relevance. Sentiment analysis revealed lower sentiment scores in participants with high Depression, Anxiety, or Agency scores, but no significant differences in Appetite or Sleep severity.

Model performance was assessed across three scenarios: text (transcripts), phone calls (audio + transcript), and video calls (audio + video + transcript). Temporal models (CNN+BiLSTM) achieved over 65% accuracy across modalities, while a fine-tuned temporal model for depression detection using video calls reached an accuracy of 81% with an f1-score of 0.79, demonstrating that our approach performs on par with state-of-the-art methods. XGBoost excelled in phone and video calls, while Ridge classifiers performed best for text-based inputs. SHAPley analysis identified key audio and video features for detecting Depression and other symptoms. A translational avatar-based interface validated system operability, demonstrating the potential for scalable, objective mental-health assessment in tele-counseling.

## INTRODUCTION

Mental health disorders represent a leading cause of disability worldwide, affecting an estimated 1 in 4 individuals over the course of their lifetime (World Health Organization [WHO], 2020). Depression, in particular, has become the leading global contributor to years lived with disability (YLD), with over 264 million people affected globally (WHO, 2017). The increasing burden of mental health conditions is exacerbated by barriers to care, including a shortage of mental health professionals, limited access to effective treatments, and the stigma surrounding mental health issues. In India, the situation is particularly urgent, with approximately 13.7% of the population suffering from some form of mental health disorder, and depression being the most prevalent, affecting around 2.7% of the population (Gururaj et al., 2016).

These statistics are compounded by the severe shortage of mental health professionals in India—there are fewer than 0.3 psychiatrists per 100,000 people, significantly below the global average (WHO, 2017; Patel et al., 2018). As a result, there is an increasing demand for scalable, efficient tools that can aid in early detection and intervention, especially in rural and underserved regions where access to trained professionals is limited. The expansion of tele-mental-health services has emerged as a promising solution to address these challenges (Ministry of Health and Family Welfare, 2020). However, the effectiveness of these services relies heavily on the availability of objective, reliable tools for assessing psychological conditions.

In tele-mental-health settings, most diagnostic processes currently depend on self-report and clinician judgment, which can often fail to capture the full complexity of psychological conditions like depression (Fitzpatrick et al., 2017). Depression itself is a highly heterogeneous condition, with patients experiencing a wide range of symptoms across cognitive, affective, physiological, and behavioral domains, such as impaired agency, sleep disturbances, appetite changes, and heightened anxiety (<u>Kroenke</u> et al., 2001). For effective intervention, it is critical to stratify depression cases based on these symptom profiles. Recent studies suggest that tailoring treatment to specific symptom clusters—such as those focused on appetite, sleep, or anxiety—can significantly enhance therapeutic outcomes (Cuijpers et al., 2020). However, mental health care systems in India often lack the resources to implement such personalized treatment approaches, and this gap in care highlights the need for automated, objective tools that can assist in the stratification and accurate diagnosis of depression.

This study aims to develop a multimodal learning framework that integrates acoustic, facial, and linguistic data to detect key psychological and behavioral conditions, including overall depression score, and subtype symptom presentations such as anxiety, appetite disturbances, agency, and sleep-related issues. By improving the stratification of depression based on symptom-dominant presentations—such as appetite, agency, anxiety, and sleep—this framework will help optimize treatment by guiding clinicians toward more personalized, targeted interventions. Ultimately, the goal is to create a scalable, reliable tool for early detection and symptom-specific intervention in tele-mental-health settings, addressing the unique challenges faced by mental health care delivery in India. Specifically, we present a multimodal learning framework for automated detection of psychological and affective attributes from behavioral interactions. We aim to integrate low-level acoustic and facial features with high-level linguistic embeddings to predict five binary outcomes: Overall Depression severity along with Appetite specific severity, Agency specific severity, Anxiety specific severity, and Sleep specific severity in symptom presentations. Our methods below comprise: (i) dataset preparation and preprocessing, (ii) modality-specific feature extraction, (iii) temporal segmentation and fusion, (iv) feature selection and reduction, and (v) supervised learning with cross-validated evaluation. Finally, we present an avatar-based interface to discuss the feasibility of the proposed tool for objective mental-health assessment in tele-counseling.

## 2. METHODOLOGY

The following section details the dataset used for model development, feature extraction from different modalities, operationalization of the output labels input feature pre-processing, model development and marginal contribution analysis.

### 2.1. Dataset description

The Extended Depression and Anxiety Interview Corpus - Wizard of Oz (EDAIC-WOZ, Gratch et al., 2014, DeVault et al., 2014) multimodal dataset containing synchronized audio, video, and text was used. The dataset was available on request to USC Institute of Creative Technologies, who gave ethical approval for this work.

The dataset consists of a wav audio file and the facial action units extracted from each frame for all the 275 participants. For each participant, archives were programmatically retrieved and decompressed to obtain raw audio and OpenFace-derived facial action unit (FAU) streams. The length of the interview ranges from 7 to 33 minutes with an average of 16 minutes.Post-traumatic Stress Disorder Checklist, Civilian Version (PCL-C) and Patient Health Questionnaire-8 (PHQ8) were collected in a self-reported manner and available for all the participants.

### 2.2. Feature extraction

Audio level and transcript level features were extracted and processed from the audio files. Video level features were available as part of the EDAIC-WOZ dataset.

#### 2.2.1. Audio level feature extraction

**eGeMAPS:** In paralinguistics analysis, the Extended Geneva Minimalistic Acoustic Parameter Set (eGeMAPS) is a widely used feature set, which we extracted using the ‘opensmil’ Python library. Initially the MP4 format of the audio data was converted to WAV format using the ‘ffmpeg’ tool to be compatible with ‘opensmil’. The audio was segmented into 30ms windows with no overlap and the feature extraction process was then performed on each window using the ‘opensmile.Smile’ class, specifying the ‘eGeMAPSv02’ feature set and ‘LowLevelDescriptors’ feature level. The extracted features consist of a set of 23 acoustic parameters.

**MFCC:** Similar to the eGeMAPS extraction method, the audio file was windowed the Mel Frequency Cepstral Coefficients were extracted using librosa library in python to end up with 13 coefficient values.

**Bag of audio words representation**: Using the 23 eGeMAPS features and 13 MFCC values, a bag of 100 audio words were constructed. K-means (K=100) clustering was performed with the features of all the 30ms windows across all the 275 participants resulting in 100 centroids. Each centroid is considered to be an audio word and the windows are assigned one of the 100 audio words.

#### 2.2.2. Text level feature extraction

Transcript generation using automatic speech recognition: Transcription used the Whisper model, producing time-aligned utterances with start/end timestamps. Segment-level transcripts were retained for synchronization with acoustic and visual features.

**Speaker Role Identification:** To differentiate the *interviewer* from the *interviewee* in these dialogues, a heuristic, multi-step algorithm was used as follows:

1. Keyword-based Identification: If a speaker mentioned the name “Ellie” (the virtual interviewer), that speaker was tagged as the *interviewer*.
2. Question Frequency Analysis: If no keyword was detected, the speaker with more questions (based on frequency of “?”) was considered the *interviewer*.
3. Tie-breaker via Verbosity: In case of equal question counts, the speaker with the shorter average utterance length was designated as the interviewer.

**Sentiment Extraction:** A transformer-based sentiment analysis model, distilbert-base-uncased-finetuned-sst-2-english from Hugging Face Transformers, was used to assess the emotional tone of each utterance. For each line spoken by the interviewee, the sentiment intensity score was computed using the following formula,

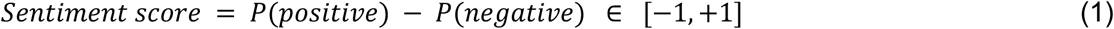

The line-level scores were averaged per participant to compute their **aggregate sentiment score**. Based on this, participants were labeled as expressing **positive**, **neutral**, or **negative** sentiment overall.

**Speaker embeddings and diarization:** Each raw audio was converted into 1-Hz voice embeddings using the VoiceEncoder module from resemblyzer library in Python. K-Means (K=2) clustering was applied to the embedding sequences to approximate two dominant speakers; segment-level speaker identity was assigned via the mode of cluster labels in a small window centered at each segment.

**Textual Feature Representation:** Utterances were grouped into overlapping text chunks of 10 utterances with an overlap of 4. Semantic embeddings were computed using the Sentence-Transformer “paraphrase-MiniLM-L6-v2” and zero padded/truncated initially to match the 87-chunk temporal resolution of the acoustic and visual streams.

#### 2.2.3. Video level feature extraction

The dataset provided the extracted facial action unit estimates for each frame restricting our potential to use any algorithms to extract additional features from the video. The mp4 file was unavailable as part of the dataset. In order to match the EDAIC-WOZ facial feature extraction method for future test data points, we developed our own pipeline based on the description provided by the authors of the dataset.

### Facial Feature Extraction

To quantitatively analyze facial behavior from the video recordings, we utilized the OpenFace 2.2.0 toolkit, a robust open-source framework for facial behavior analysis. OpenFace processes video streams on a frame-by-frame basis to identify and track facial landmarks, from which it derives a comprehensive set of facial features.

For our study, we configured the toolkit to extract three specific sets of features from each video file (.mp4) as explained below:

**Facial Action Units (FAUs**): We extracted the intensity and presence of 17 AUs based on the Facial Action Coding System (FACS). These FAUs represent the fundamental muscle movements of the face (e.g., AU12 for Lip Corner Puller, associated with smiling). The output gives us a continuous value for the intensity of each AU per frame.

**Head Pose**: The 3D orientation and positioning of the head were extracted for each frame. This is represented by six variables: three for translational movement in millimeters (T_x, T_y, T_z) and the other three for rotational movement in radians (pitch, yaw, roll). This data allows for the normalization of facial attributes and the analysis of head gestures.

**Eye Gaze**: The direction of eye gaze was captured as 3D gaze vectors for both the left and right eyes. This provides detailed information on the subject’s point of attention throughout the recording.

The extraction process was automated via the command-line interface of OpenFace. The toolkit generated a time-stamped, frame-by-frame output in a comma-separated values (CSV) format. Each row in the resulting CSV file corresponds to a single video frame, and the columns contain the timestamp, frame number, and the full set of extracted FAU, head pose, and eye gaze features. This structured time-series data was used for our subsequent analysis and model testing stage.

### 2.3. Cross modality alignments - input generation

**Multimodal Fusion:** For each participant, the features were averaged for a 30 second window with 50% overlap. For textual features, utterances were grouped into overlapping text chunks of 10 utterances with an overlap of 4. The time-aligned chunk representations from audio, text, and visual modalities were concatenated along the feature axis to form a unified multimodal tensor: a maximum length sequence of 87 chunks (corresponding to mean duration), each containing fused acoustic, semantic, and facial features. Shorter sequences were zero-padded and longer sequences were truncated in the front. This served as input to downstream models (Nykoniuk, 2025).

**Summary level features:** The above-mentioned time-series data was used to train temporal models. In order to train simple Machine Learning models for explainable purposes, summary level features such as mean, range and standard deviation for each feature were extracted and used as input.

### 2.4. Output operationalization/Ground-Truth Generation

In the study, *ground-truth outcomes* refer to the reference labels used to train, validate, and evaluate predictive models based on the EDAIC dataset. Because the analysis targeted multiple psychological and behavioural domains, it was necessary to construct consistent and clinically interpretable binary variables representing each condition.

Five domains measured within the dataset were examined:

1. Overall Depression score,
2. Appetite-related disturbances,
3. Agency (perceived control or self-efficacy),
4. Anxiety, and
5. Sleep-related disorders.

All labels were derived according to validated scoring rules and threshold criteria defined in the *EDAIC Data Manual*, ensuring reproducibility and alignment with established psychometric standards.

**Depression**: Depressive status was determined from PHQ-8 totals. Scores > 10 indicated 1 (participant exhibiting depressive symptoms), while scores ≤ 10 indicated 0 (participant having no/mild depressive symptoms). This threshold corresponds to established PHQ-8 conventions for clinically significant depression as provided in the EDAIC Data manual.

**Appetite:** Appetite disturbance was derived from the PHQ-8 item: “Over the last two weeks, how often have you been bothered by poor appetite or overeating?” (PHQ8_5_Appetite). The PHQ-8 instrument specifies this item on a 4-point scale (0 = Not at all, 1 = Several days, 2 = More than half the days, 3 = Nearly every day) (Spitzer, 1999, Kroenke, 2001). This item has been validated as part of the PHQ-8 in both general and clinical populations (Kroenke, 2009, 2017; Pavlov, 2022). While prior studies have primarily used the total PHQ-8 score for overall depression severity, the current analysis adopts a domain-specific operationalisation focused on the appetite item itself. Responses ≥ 2 (more than half the days or nearly every day) were labeled 1 (participant exhibiting appetite issues), and responses ≤ 1 were labeled 0 (participant not exhibiting appetite issues). This dichotomisation captures clinically meaningful appetite-related disturbance.

**Agency:** Psychological agency was computed from both PHQ-8 and PCL-C items reflecting “motivation”, “mood”, and “self-control”. PHQ-8 items on “loss of interest”, “depressed mood”, “feelings of failure”, and “concentration difficulties” were reverse-scored to represent higher agency. Corresponding PCL-C items covered cognitive/mood domains (emotional detachment, numbness, a foreshortened future) and hyperarousal domains (vigilance, difficulty concentrating) to capture diminished cognitive regulation and heightened arousal. The two composite scores were averaged to obtain an overall Agency Score, which was dichotomised at the sample median (≥ median → label 1: participant exhibiting higher agency; < median → label 0: participant exhibiting lower agency).

This operationalization is grounded in the conceptualization of agency as the capacity for emotion regulation and cognitive regulation (Daros, 2021; Megreya, 2025) and aligns with evidence linking self-regulatory deficits (e.g., reduced motivation, concentration difficulties, hyperarousal) with poorer psychological outcomes (Ohata, 2020).

**Anxiety:** Anxiety was measured through nine items capturing somatic and hyperarousal symptoms: four from the PHQ-8 (“sleep”, “tired”, “concentrating”, “psychomotor agitation”) and five from the PCL-C (“sleep”, “irritability”, “concentrating”, “hyperalertness”, “jumpiness response”). For each participant, responses were summed to compute an overall Anxiety Score, which was dichotomised at the sample median into a binary label: **1** *(participant exhibiting higher anxiety)* and **0** (*participant exhibiting lower anxiety)*.

This operationalization reflects the established conceptualization of anxiety as a combination of somatic (fatigue, sleep) and hyperarousal (vigilance, jumpiness) symptoms (Peters, 2021; Peng, 2022; Dong, 2025), and leverages a median-split approach that has been used in other symptom-based studies of anxiety and depression (Peng, 2022).

**Sleep-Related Disorders**: Sleep disturbance was assessed using two items PHQ8_3 (*trouble falling or staying asleep / sleeping too much*) and PCL-C_13 (*trouble falling or staying asleep*). Their combined score was dichotomized at the median, yielding 1 *(participant exhibiting significant sleep disturbance)* and 0 *(participant without significant sleep disturbance)*.

### 2.4. Feature pre-processing

#### 2.4.1. Information-Theoretic Screening and Redundancy Control

Feature curation proceeded per label in three stages. First, mutual information (MI) between each feature and the target was estimated to capture nonlinear dependencies; features with zero MI were removed as non-informative. Second, pairwise correlations were computed within the MI-retained set and highly collinear pairs (absolute Pearson correlation exceeding 0.8) were pruned by retaining the feature with the higher MI score, thereby reducing redundancy and variance inflation. Third, the top 20% of features by MI rank were selected to yield a compact, task-specific subset that respects differences in what drives each outcome.

#### 2.4.2. Standardization and Class Rebalancing

All features were standardized to zero mean and unit variance prior to modeling to place heterogeneous predictors on a comparable scale. To mitigate class imbalance without contaminating evaluation, synthetic minority over-sampling (SMOTE) was applied strictly within the training partitions for each label. This procedure improved minority-class sensitivity while preserving the original distribution in validation folds.

#### 2.4.3. Quality Controls and Robustness

Variance and single-level filtering preceded MI to remove degenerate predictors. Correlation pruning followed MI so that retained features remained informative yet minimally redundant, reducing overfitting risk. Standardization and resampling were confined to training folds to prevent target leakage. All steps were executed independently per label, ensuring that feature subsets and explanations reflect label-specific signals rather than a shared compromise across tasks.

#### 2.4.4. Handling Sequence Length Variability

To avoid learning from padding artifacts, zero-padding was removed prior to model input, and variable-length (ragged) sequences were supported. During batching, sequences were converted to dense tensors after trimming to preserve authentic temporal structure.

### 2.5. Model development

#### 2.5.1. Model Architecture

A hybrid Convolutional–Bidirectional LSTM (CNN–BiLSTM) classifier modeled local and long-range temporal dependencies. The network comprised a 1D convolution (64 filters; kernel size 3; ReLU), batch normalization, dropout (0.3), a bidirectional LSTM (64 units), global max pooling, two dense layers (32 and 16 units; ReLU), and a sigmoid (or softmax) output for binary prediction. Optimization used Adam (learning rate 0.001) with binary cross-entropy and a step-down learning-rate schedule after 20 and 40 epochs. Training ran for 15 epochs with batch size 16. Additionally, various simple Machine Learning models were trained and tested on summary level features as reported in the table 2 of results section.

**Table 1:** Univariate statistical analysis of aggregate sentiment scores. The table presents sentiments based predictions of several subtypes of depression, and is overall severity scores. Based on normality of the classes for various labels, the test used differed. T represents True and F represents False for normality test.

| Label | Sample size<br>(classes 0, 1) | Normality<br>(classes 0, 1) | Test used | Mean sentiment<br>(classes 0, 1) | p value |
| --- | --- | --- | --- | --- | --- |
| **Depression** | **209, 66** | **T, F** | **Mann Whitney U** | **0.079, 0.006** | **0.01** |
| Appetite | 198, 77 | T, F | Mann Whitney U | 0.074, 0.029 | 0.05388 |
| **Agency** | **136, 139** | **F, T** | **Mann Whitney U** | **0.002, 0.120** | **0.00001** |
| **Anxiety** | **126, 149** | **T, T** | **t-test** | **0.117, 0.015** | **0.00028** |
| Sleep | 175, 100 | T, T | t-test | 0.076, 0.036 | 0.18195 |

**Table 2:** Model performance across 3 scenarios. The accuracy (sensitivity, specificity) has been reported for various models and feature types. Based on the geometric mean of accuracy, sensitivity and specificity, the models have been ranked for each scenario and labeled with green corresponding to the best model and red corresponding to the worst model.

| TRANSCRIPT |  |  |  |  |  |  |
| --- | --- | --- | --- | --- | --- | --- |
| Feature | Model | Appetite | Anxiety | Sleep | Agency | Depression |
| Summary | *Logistic Reg* | 53 (51, 53) | 60 (69, 50) | 57 (52, 61) | 55 (60, 51) | 54 (51, 54) |
|  | *MLP* | 52 (46, 54) | 56 (73, 37) | 59 (56, 61) | 49 (38, 60) | 60 (49, 63) |
|  | *XGBoost* | 56 (30, 65) | 59 (64, 52) | 54 (40, 62) | 55 (55, 54) | 63 (30, 74) |
|  | *SVM* | 51 (55, 49) | 58 (73, 41) | 55 (41, 63) | 52 (59, 45) | 51 (42, 53) |
|  | *Ridge* | 53 (52, 53) | 61 (69, 51) | 58 (53, 62) | 56 (61, 51) | 54 (55, 54) |
| Temporal | *CNN+BiLSTM* | 66 (48, 73) | 66 (67, 65) | 62 (56, 65) | 62 (66, 57) | 65 (38, 73) |
| TRANSCRIPT + AUDIO |  |  |  |  |  |  |
| Feature | Model | Appetite | Anxiety | Sleep | Agency | Depression |
| Summary | *Logistic Reg* | 57 (49, 60) | 61 (60, 62) | 64 (62, 65) | 60 (61, 60) | 62 (50, 65) |
|  | *MLP* | 52 (46, 54) | 49 (51, 47) | 51 (48, 53) | 49 (51, 47) | 54 (41, 59) |
|  | *XGBoost* | 60 (54, 62) | 62 (63, 62) | 61 (61, 61) | 61 (66, 56) | 66 (53, 70) |
|  | *SVM* | 50 (53, 48) | 44 (30, 61) | 46 (50, 44) | 47 (43, 50) | 45 (55, 41) |
|  | *Ridge* | 59 (47, 63) | 59 (59, 59) | 64 (61, 65) | 61 (62, 61) | 65 (51, 70) |
| **Temporal** | *CNN+BiLSTM* | 63 (49, 68) | 59 (63, 54) | 65 (69, 63) | 63 (68, 58) | 72 (49, 80) |
| **TRANSCRIPT + AUDIO + VIDEO** |  |  |  |  |  |  |
| **Feature** | **Model** | **Appetite** | **Anxiety** | **Sleep** | **Agency** | **Depression** |
| **Summary** | *Logistic Reg* | 56 (60, 54) | 56 (76, 32) | 56 (69, 49) | 57 (70, 43) | 62 (61, 62) |
|  | *MLP* | 60 (51, 64) | 56 (64, 47) | 62 (63, 61) | 60 (67, 52) | 67 (50, 72) |
|  | *XGBoost* | 60 (43, 67) | 54 (73, 32) | 61 (57, 63) | 57 (70, 43) | 68 (44, 76) |
|  | *SVM* | 65 (47, 72) | 55 (97, 5) | 64 (70, 61) | 50 (99, 1) | 73 (44, 82) |
|  | *Ridge* | 38 (85, 19) | 56 (95, 10) | 48 (92, 23) | 51 (91, 10) | 43 (89, 28) |
| **Temporal** | *CNN+BiLSTM* | 73 (43, 85) | 64 (65, 64) | 69 (56, 74) | 71 (70, 71) | 81 (87, 78) |

#### 2.5.2. Evaluation Protocol

A stratified 10-fold cross-validation procedure was used to obtain a robust estimate of model performance, ensuring that each fold preserved the original class distribution. Performance was evaluated by averaging metrics across all test folds, including accuracy, the overall proportion of correct predictions; sensitivity (recall), measuring the ability to correctly identify positive cases (TP / (TP + FN)); and specificity, reflecting the ability to correctly identify negative cases (TN / (TN + FP)).

#### 2.5.3. Fine-tuned Model for Depression Label prediction

In order to match the methodology adopted by the state-of-the-art models on EDAIC dataset analysis, we performed holdout validation on the provided split in the dataset. As we observed that the temporal model yielded the best metric, we developed a fine-tuned CNN-BiLSTM model with Attention layers to predict depression labels. Data augmentation and class balancing was performed by randomly cropping 75% of the timeseries datapoint, merging randomly selected two of the 35% chunks of same subject, adding gaussian noise jitters, random feature dropout, masking contiguous time slices with zeros and randomly scaling by a small factor. The ensured that the model does not overfit the small training dataset. All the time-series data were resampled to have 37 timesteps (equivalent to 10 minutes based on section 2.3).

The model architecture is presented in figure 1. The model was trained using the class balanced version of focal loss with gamma value as 1.25. Adam optimizer was used with learning rate of 0.0005, batch size of 8 and number of training epochs as 20.

**Figure 1:**
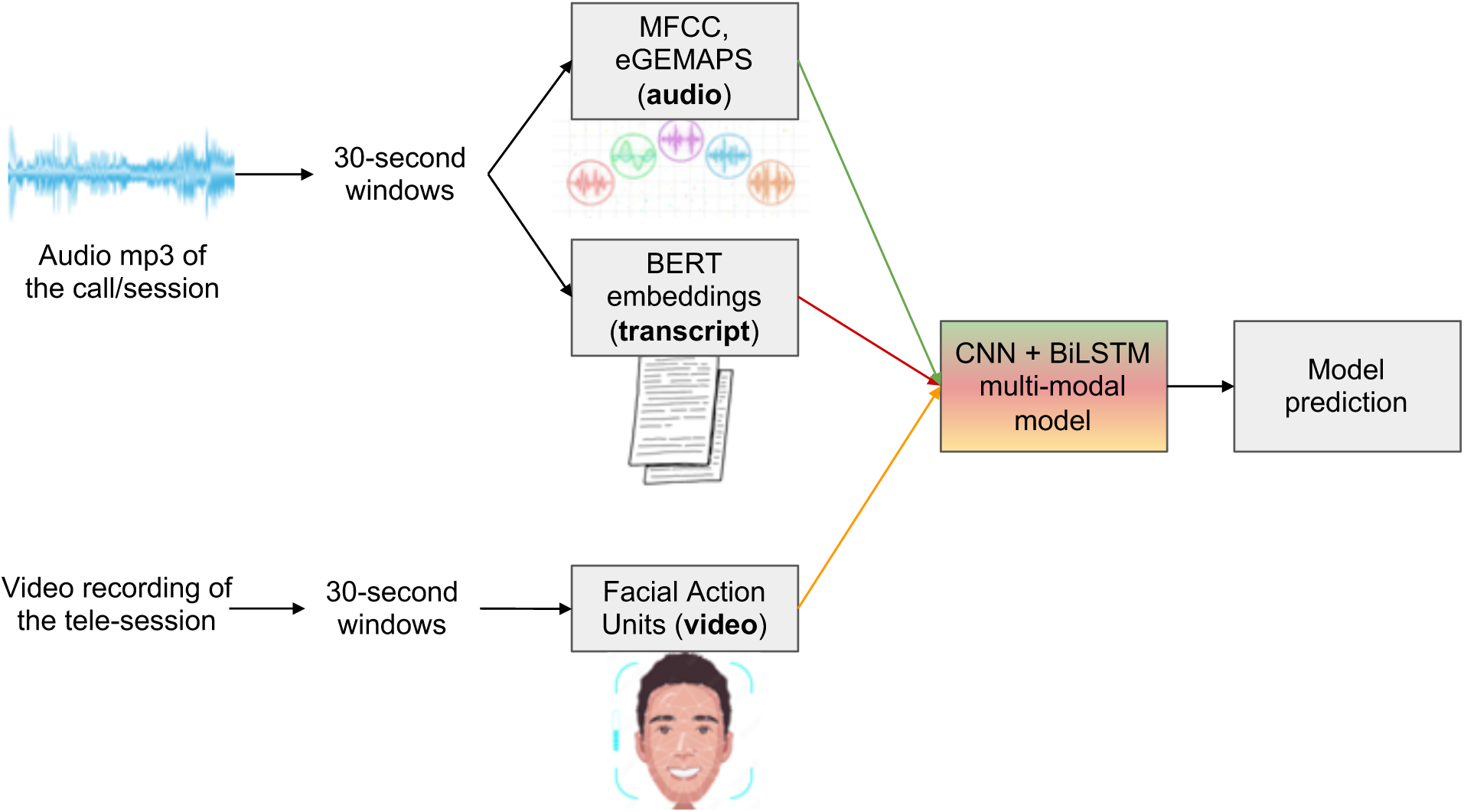
Overview of the prediction model pipeline. Based on the available input modality, the multi-modal model is deployed for prediction of the output labels. The detailed architecture of the deep network model is provided in the inset.

### 2.6. Feature importance

#### 2.6.1. Predictive Modeling for Explainability

For explainability-oriented prediction, gradient-boosted decision trees (XGBoost) were trained separately for each label using a binary logistic objective with log-loss monitoring. Boosted trees were chosen for their strong performance on mixed-scale tabular data and their compatibility with TreeSHAP, which provides faithful local and global attributions. While sequence models are used elsewhere in the pipeline, tree models here serve as a transparent lens on feature contributions.

#### 2.6.2. SHAP Explainability and Category-Level Attribution

Shapley values were computed using TreeSHAP and averaged in absolute value across participants to obtain global feature importance. Two complementary summaries were constructed. First, OpenFace features were aggregated by anatomical category (Eyes, Brows, Lips/Mouth/Jaw, Nose, HeadPose), revealing region-level drivers of each label; because each descriptor appears with multiple statistics, category totals reflect both steady levels and variability or extremes. Second, acoustic descriptors were summarized by family, separating MFCC from eGeMAPS to distinguish spectral-envelope cues from prosodic and voice-quality characteristics. For fine-grained interpretation, individual features were ranked within each category and numeric OpenFace identifiers in the rankings were resolved to semantic names (e.g., AU12; lip corner puller_max).

#### 2.6.3. Integrated gradient explainability of temporal model

The same categories as above were used for explaining the aggregated feature importance score across categories and timesteps. Integrated gradient for all test datapoints belonging to class 1 from zero vector baseline as input was computed and averaged across features belonging to each category to obtain the category level feature importance score.

#### 2.6.4. Sentiment analysis

We examined whether participants’ aggregate sentiment scores differed significantly across six binary clinical labels: Overall Depression, and specifically Appetite, Agency, Anxiety, and Sleep dominant symptoms. For each label, participants were divided into two groups based on their classification (Group 0 = label 0, Group 1 = label 1). The distribution of sentiment scores within each group was first evaluated using the Anderson–Darling test for normality. Depending on the outcome, two types of inferential tests were applied: Welch’s *t*-test when both groups met normality assumptions, or the non-parametric Mann–Whitney U test when either group violated them. Statistical significance was defined as *p* < 0.05. To contextualize results, we additionally flagged comparisons exhibiting substantial class imbalance, defined as any group containing fewer than 40% of the samples in the other group.

### 2.7. Translational Application Design and Pilot Feasibility

As a translational outcome of this study, we propose an avatar-based tele-counseling application designed to operationalize multimodal behavioral assessment in privacy-sensitive settings. The system enables real-time capture of facial action units (FAUs) via webcam using Google MediaPipe, which are mapped onto a virtual avatar to preserve expressive fidelity while anonymizing user identity. Frame-wise FAU features are logged for downstream multimodal analysis.

In parallel, audio streams are transcribed using a third-party speech-to-text service that performs speaker diarization, language detection, and timestamped transcription, enabling alignment between visual, acoustic, and linguistic modalities. A primitive pilot deployment was conducted to validate system functionality, data quality, and end-to-end operability. The pretrained model predictions were not surfaced to counselors or used to inform therapeutic decisions, serving instead to verify the feasibility of integrating multimodal prediction within live tele-counseling workflows.

## RESULTS

### 3.1. Dataset summary

The files from all 275 participants were processed and features were extracted. Post dichotomization of the output labels, we observe class imbalance towards class 1 (no issues) for Depression label because the dataset was primarily collected from psychotypical individuals. Agency and Anxiety labels are balanced because of median split (Please see Supplementary table 1, figure 1 for more details on class labeling). The distribution in class 0 (low scores) and class 1 (high scores) is shown in Supplementary Figure 2. It is to be noted that participants belonging to class 0 are healthier than those belonging to class 1 except agency labels.

### 3.2. Sentiment analysis

Participants with high overall **Depression score, and specifically Agency, or Anxiety** symptoms exhibited significantly lower aggregate sentiment scores compared to their counterparts, suggesting that these clinical conditions are associated with detectable negative emotional tone in speech. In contrast, the Appetite and Sleep labels did not show statistically significant differences in sentiment, which may reflect the subtler nature of these symptoms or potential noise and inconsistency in the corresponding labels.

### 3.3. Model performances across scenarios

In order to mimic various real-life scenarios, three different combinations of input features were considered for model development. In telehealth, texts, phone calls and video calls are most commonly used for conversing with the clinician. Considering those scenarios, transcript level features are used for mimicking texts, transcript and audio level features for phone calls and all 3 modalities (transcript, audio, video) were used for video calls. Summary feature models and time-series feature models for all the labels across 3 scenarios were developed and compared.

From table 2, we notice that across different scenarios, models trained on temporal features are able to perform well above chance level for all the labels highlighting the presence of a dynamic pattern in detecting psychological distress. The models trained on temporal inputs are able to perform with an accuracy level of >65% for rich modality inputs across all labels, while the other models are inconsistent depending on the modality and label.

In case of summary features, for transcript-level inputs, Ridge classifier works the best for Appetite, Anxiety and Agency, MLP works best for Sleep and Depression. Overall, Ridge classifier is the best model for detecting psychological issues in a text based scenario.

When we are dealing with phone and video calls, XGBoost ranks the best across all labels and scenarios and Logistic Regression ranks second. Therefore, as the XGBoost classifier worked best with rich modalities, the same was used for SHAPley analysis in the next section.

### 3.4. Fine-tuned model performance on predicting depression label

As compared to the state-of-the-art model for predicting depression labels (Lin Lin, 2020), our architecture and approach is robust, subject-wise predictive and uses a relatively simplistic model with way fewer parameters that has deployable ability for real-time purposes. Across 5 iterations, we obtained an average F1 score of 0.79, accuracy of 81%, sensitivity 87% and specificity 78% on the holdout dataset.

### 3.5. Feature importance using SHAPley

SHAPley values highlight the marginal contribution of each feature towards the prediction of a class. In order to make the explanation language agnostic, only audio and video features were explained through SHAPley analysis. The sum of absolute SHAPley values of the features in a category for a datapoint represents the marginal contribution of a category towards the prediction. In Figure 2, the mean of marginal contributions of each category across datapoints are presented. As there were more audio features, the value is higher for audio feature categories.

**Figure 2:**
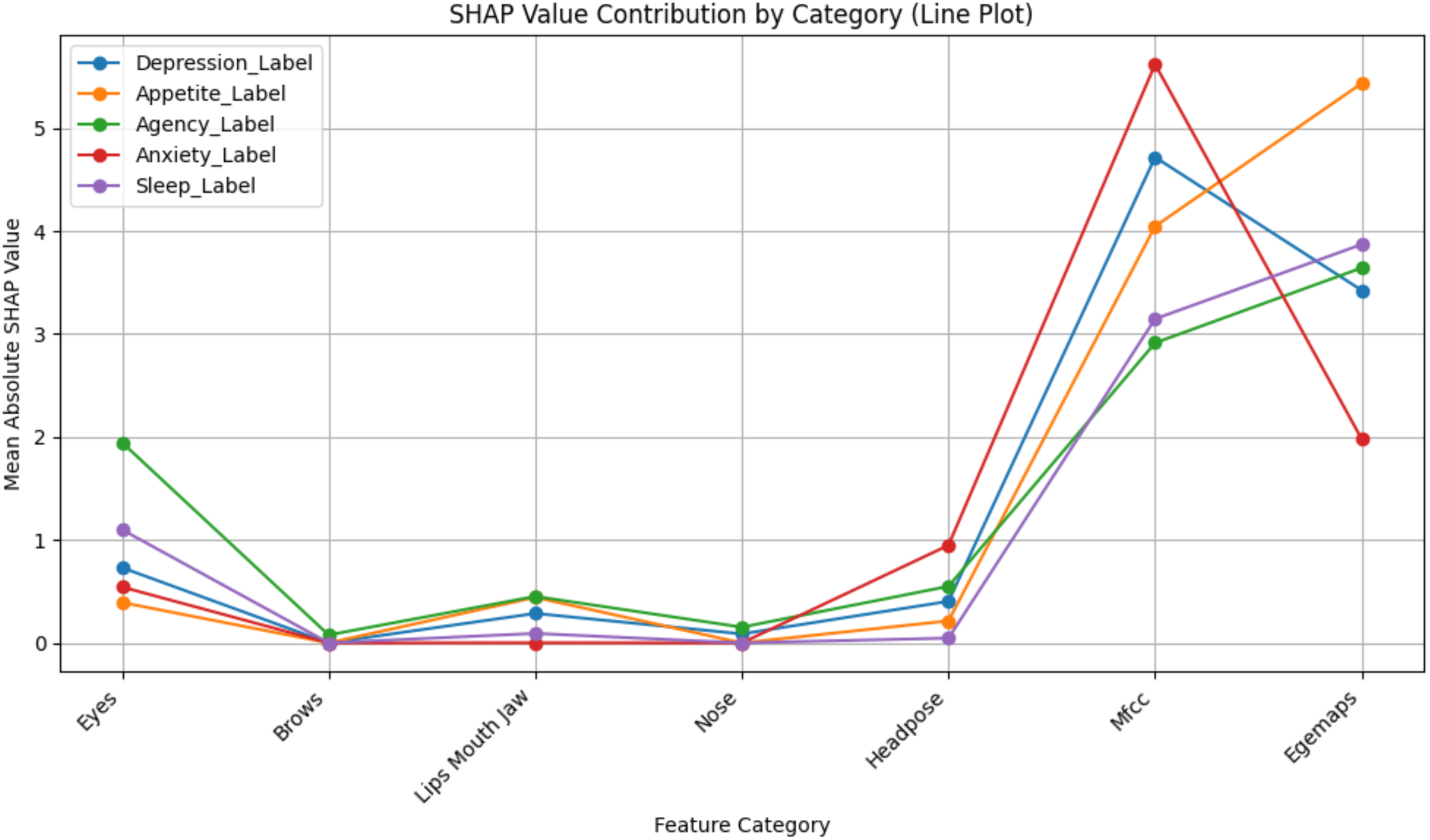
Marginal contribution of features in each category towards prediction of all labels. The figure presents marginal contributions in terms of mean absolute SHAP values of specific facial and audio features for predicting depression severity and its subtype severity.

From figure 2, we can observe that Eyes are sensitive for Agency, Jaw for Appetite, headpose for Anxiety, MFCC for Anxiety, Agency, Depression and Appetite and EgeMAPS for Appetite. Audio features are able to detect the nuances of Depression and Appetite issues while video features are unable to detect Depression. Eyes are able to detect sleep and agency issues.

From Figure 3, we observe that depressed participants showed several distinctive facial and head-movement patterns compared to healthy individuals. Their left-eye gaze was more unstable, as indicated by greater depth-wise standard deviation, and they exhibited the greatest sideways head movement overall. For the right eye, depressed participants tended to gaze lower, reflected in lower minimum values, and they also showed a reduced smile extent. They displayed less nose wrinkling and did not move their heads closer to the camera, as shown by lower maximum head-position values. Additionally, their right eye sat lower relative to healthy participants, and their average sideways head position tended to be more leftward.

**Figure 3:**
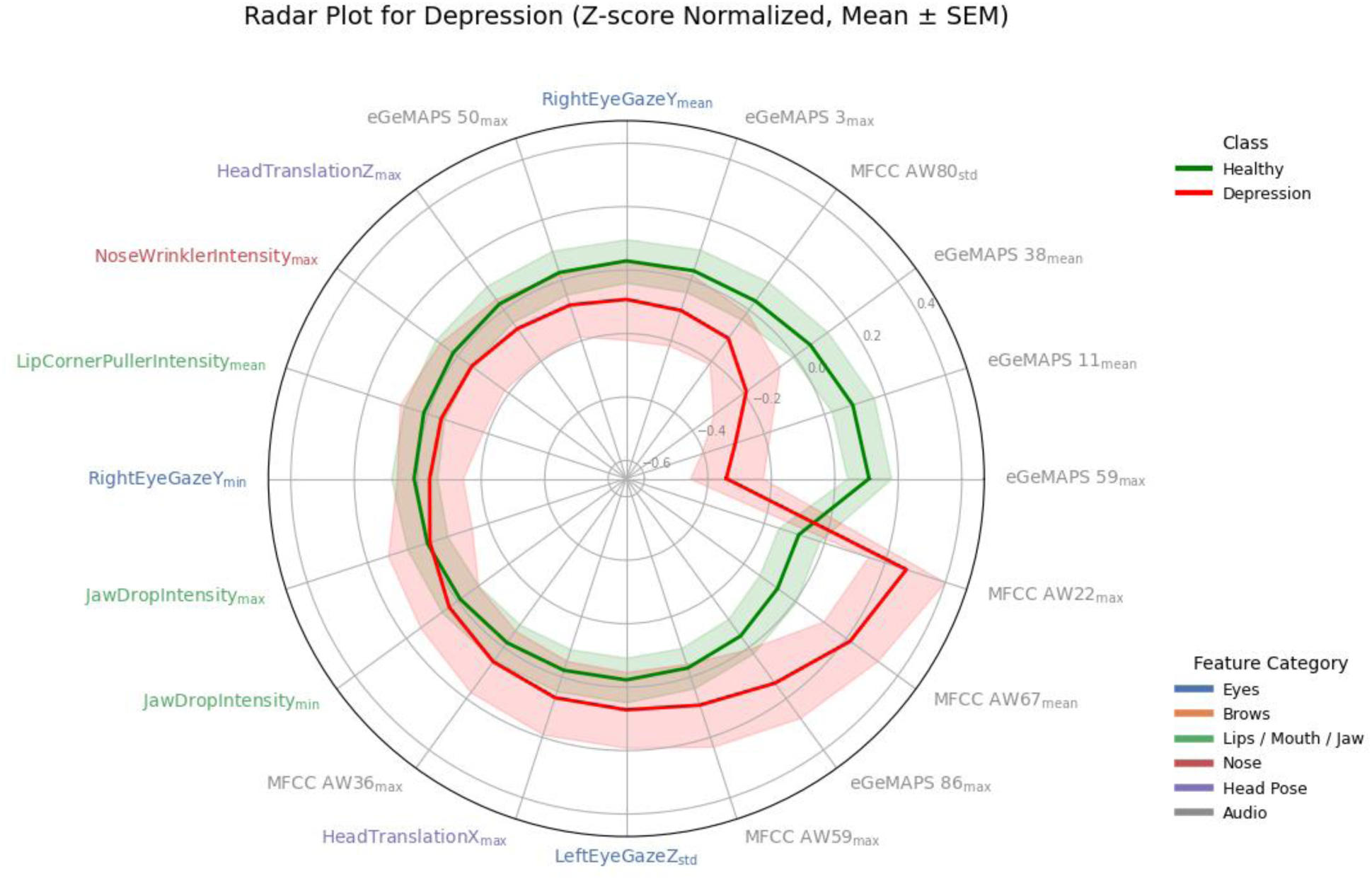
Features of importance for predicting Depression. The normalized values of top features contributing towards the prediction of Depression label were plotted as radar plot. AW for audio features correspond to audio words and subscript text represent the summary feature. The features are positioned from most to least contributing towards healthy class in anticlockwise direction.

### 3.6. System Evaluation and Practical Feasibility

A preliminary pilot implementation was conducted to assess the operational feasibility of deploying the proposed multimodal assessment framework within an avatar-based tele-counseling environment. The system successfully captured synchronized facial action unit (FAU), acoustic, and linguistic data streams during live counseling sessions without interrupting therapist–client interaction. Frame-wise FAU extraction and logging operated continuously, and audio transcription with speaker differentiation and timestamp alignment was completed for all pilot sessions. The pilot confirmed end-to-end system operability, including real-time data acquisition, multimodal alignment, and storage for downstream analysis. No session-level failures or workflow disruptions were observed, indicating that the proposed application design can support future controlled evaluations of multimodal behavioral modeling in real-world tele-mental-health settings.

The automated PHQ-9 scoring module demonstrated the feasibility of generating interpretable, timestamp-aligned symptom estimates directly from conversational data, supporting rapid post-session assessment. Together, these test applications validate the practical viability of deploying multimodal behavioral models within real-world tele-mental-health workflows.

## DISCUSSION

In this study, we introduced a multimodal learning framework to automatically detect psychological and affective attributes from behavioral interactions. Our approach integrates acoustic, facial, and linguistic features to predict five psychological conditions: Overall Depression, Appetite-related disturbances, Agency, Anxiety, and Sleep disturbances. The results highlight the potential of multimodal systems to support objective and efficient mental health assessments, especially in contexts where traditional diagnostic resources are scarce.

### Key Findings and Their Implications

Our findings indicate that the Ridge Classifier outperformed Logistic Regression in terms of specificity for labels such as Overall Depression, Appetite disturbances, and Sleep disorders, prioritizing the reduction of false positives. This aligns with previous research which emphasizes the importance of specificity, particularly in conditions with complex symptomatology and overlapping diagnostic features (Cuijpers et al., 2020; Hussain et al., 2020). Conversely, Logistic Regression showed higher sensitivity for Anxiety, identifying more positive cases at the cost of increased false positives, suggesting that different classifiers may be better suited for different diagnostic objectives. Overall, across multimodals, XGBoost classifier ranks the best based on the geometric mean of accuracy, sensitivity and specificity across labels.

One important advantage of our system is its flexibility in using different modalities. While the full multimodal framework, incorporating audio, video, and text, provides the most comprehensive analysis with the prediction accuracy 81% for depression label using the fine-tuned temporal model, the system is capable of functioning reasonably well with just a subset of these modalities. For example, the system can still perform at a decent accuracy above chance level for predicting depression using audio-only (72%) or text-only data (65%), or combinations of two modalities i.e., audio + text (72%). This adaptability is crucial, particularly in real-world settings where some modalities may be unavailable or impractical to collect. In practice, being able to rely on just one modality, like audio for detecting depression or anxiety through speech patterns, or video for detecting facial expressions associated with traumatic stress disorder (labeled in the dataset), provides a significant advantage in scalability and accessibility (Zeng et al., 2007).

Another feature of our model is the interpretability of the utilized machine learning models. Model transparency is essential, particularly in clinical settings where clinicians need to understand the reasoning behind the system’s predictions to build trust and ensure its effective application in practice (Lipton, 2018).

### Positioning our study relative to prior work

State of the art models on EDAIC-WOZ dataset used in our study have relatively heavier architecture with the number of trainable parameters is way larger than the datapoints available to predict depression severity, and has reported an f1 score as high as 0.85. For example, in a study (Lin, 2020), the authors used CNN-BiLSTM model like our study, resampled the datapoints into unnaturally larger segments (about 15secs) for training and validation purposes and reported an f1score of 0.85. In another recent study (Nykoniuk, M, 2025), the authors developed a fusion Bi-LSTM model with Attention and reported an f1 score of 0.79 upon validation.

### Limitations and Future Work

Despite the promising results, several limitations must be addressed. First, while the EDAIC dataset is rich in multimodal data, it may not fully capture the diverse manifestations of psychological disorders across different cultural contexts. Given that mental health symptoms can vary significantly due to cultural, social, and environmental factors, further research should explore the generalizability of our model to other populations (Gururaj et al., 2016). Future work should incorporate more diverse and larger datasets to enhance model robustness, and also fine tuning based on region-specific datasets. Secondly, the EDAIC dataset has only ∼10% of the population as clinically diagnosed with depression. Therefore, the model requires more data points from clinically depressed populations for robust and reliable deployment in real-world settings.

To conclude with, through the above study, we tested the feasibility of developing an assistive tool such as multi-modal models for predicting various attributes of psychological disorders that can help the clinician in developing a summarized report for elevating the quality of mental help through tele-sessions.

## Data Availability

No new data is generated in the study.

## ACKNOWLEDGEMENTS

The authors thank for the fruitful discussions with Pushkar and Ashutosh Modi from IIT Kanpur, Sangath NGO in India for allowing us to pilot test the interface.

## AUTHOR CONTRIBUTIONS

PPB, AJAF and LT contributed to the conceptualization, writing of the manuscript. PPB, AJAF prepared the final draft of the manuscript, AJAF, AR, NP, RG, VK, VK contribute to the data analysis and writing of the manuscript, AT, AS, OM, KJ contributed to study experimentation

## DATA AVAILABILITY STATEMENT

No new data is generated in the study.

## SUPPLEMENTARY MATERIAL

The detailed mathematical formulas for each scoring and labelling are given in **Figure 1**. The specific mapping of questionnaire items to their associated psychological and behavioral domains is summarized in **Table 1**.

**Figure 1.**
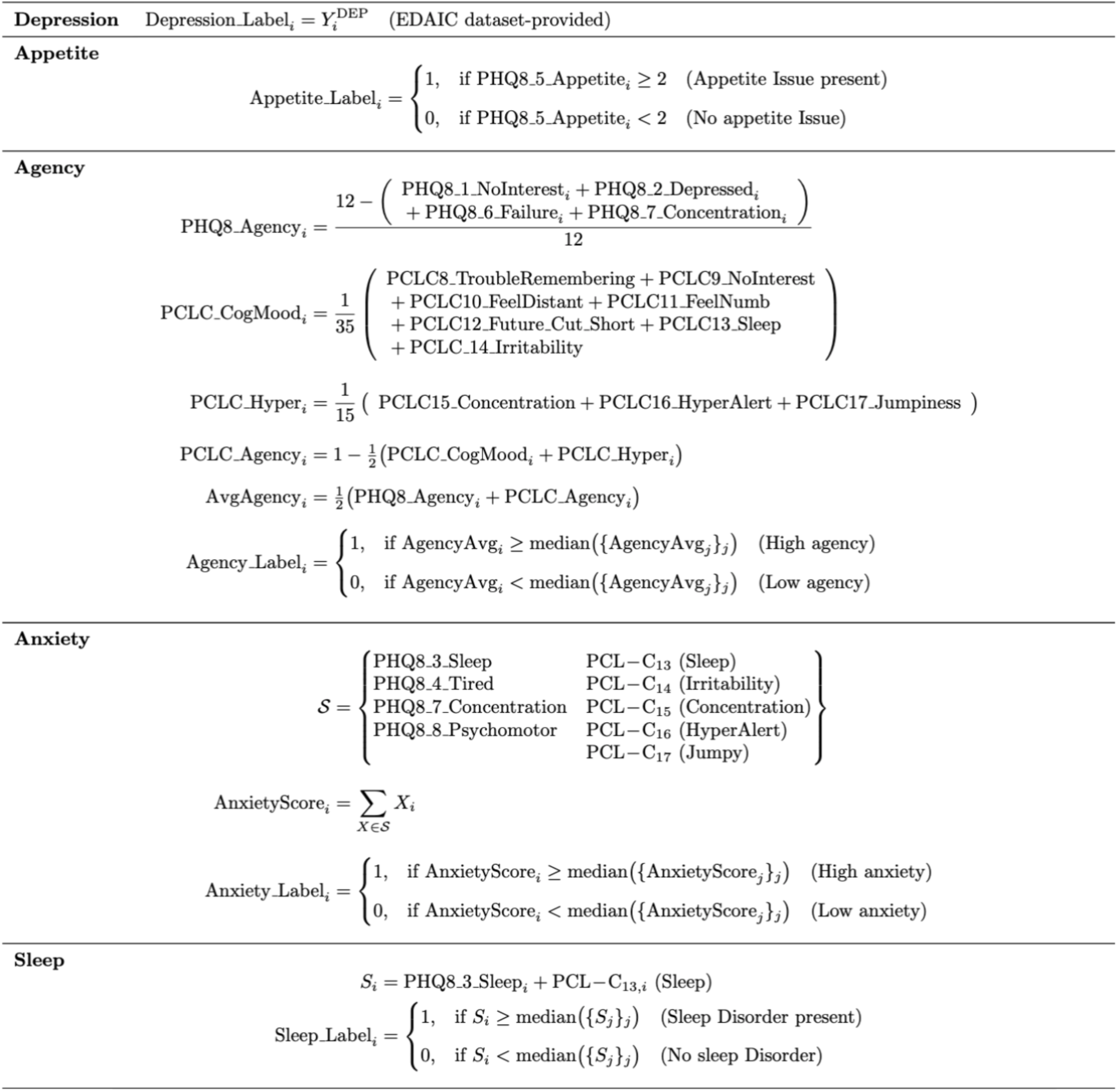
Mathematical Formulas used to generate binary outcome labels for Overall Depression, Appetite, Agency, Anxiety and Sleep disorders. *subscript *‘i’* indexes participants.

**Figure 2:**
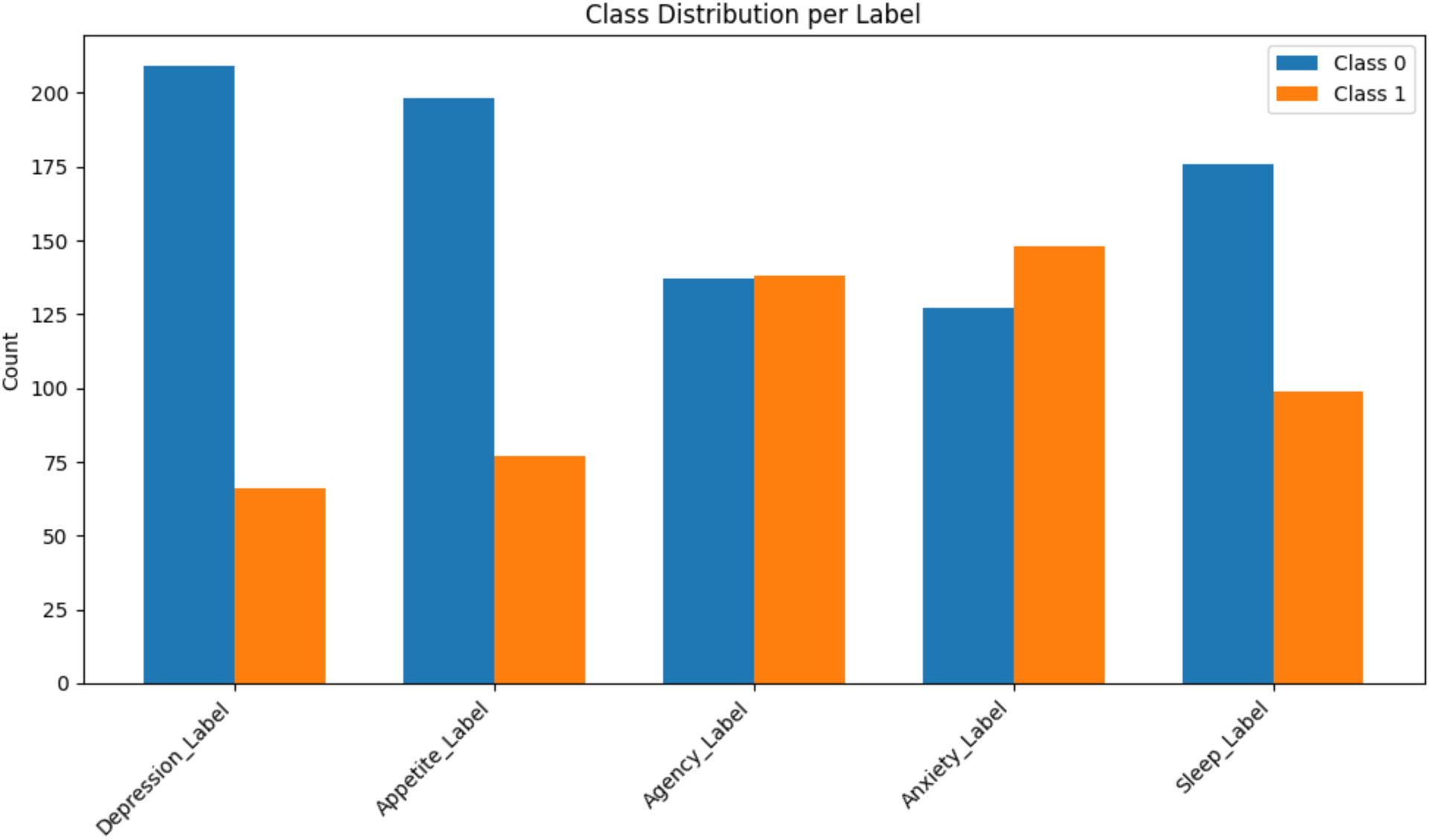
Distribution of participants for each class label.

**Table 1.** Mapping of questionnaire items to psychological and behavioral domains. summarizes the mapping between individual items from the PHQ-8 and PCL-C instruments and the six psychological and behavioral domains examined in this study: Depression, Appetite, Agency, Anxiety, and Sleep-related disorders. Each questionnaire item was aligned with one or more domains based on its underlying construct. This mapping guided the generation of domain-specific ground-truth labels for subsequent modeling and analysis.

| Item Name | Question / Construct | Associated Domain |
| --- | --- | --- |
| PHQ8_1_NoInterest | "Little interest or pleasure in doing things" | Depression, Agency |
| PHQ8_2_Depressed | "Feeling down, depressed, or hopeless" | Depression, Agency |
| PHQ8_3_Sleep | "Trouble falling or staying asleep, or sleeping too much" | Anxiety, Sleep |
| PHQ8_4_Tired | "Feeling tired or having little energy" | Anxiety |
| PHQ8_5_Appetite | "Poor appetite or overeating" | Appetite |
| PHQ8_6_Failure | "Feeling bad about yourself — that you are a failure or have let yourself or your family down" | Agency |
| PHQ8_7_Concentration | "Trouble concentrating on things" | Agency, Anxiety |
| PHQ8_8_Psychomotor | "Moving or speaking so slowly that other people could have noticed, or being fidgety or restless" | Anxiety |
| PCL-C_8_TroubleRemembering | "Trouble remembering important parts of the stressful experience" | Agency |
| PCL-C_9_NoInterest | "Loss of interest in activities you used to enjoy" | Agency |
| PCL-C_10_FeelDistant | "Feeling distant or cut off from other people" | Agency |
| PCL-C_11_FeelNumb | "Feeling emotionally numb or unable to have loving feelings" | Agency |
| PCL-C_12_Future_Cut_Short | "Feeling as if your future will somehow be cut short" | Agency |
| PCL-C_13_Sleep | "Trouble falling or staying asleep" | Anxiety, Sleep |
| PCL-C_14_Irritability | "Feeling irritable or having angry outbursts" | Anxiety |
| PCL-C_15_Concentration | "Having difficulty concentrating" | Agency, Anxiety |
| PCL-C_16_HyperAlert | "Being 'super-alert' or watchful on guard" | Anxiety |
| PCL-C_17_Jumpy | "Feeling jumpy or easily startled" | Anxiety |
| Depression_label | Binary depression label | Depression |

## REFERENCES

1. Gratch J, Artstein R, Lucas GM, Stratou G, Scherer S, Nazarian A, Wood R, Boberg J, DeVault D, Marsella S, Traum DR. The Distress Analysis Interview Corpus of human and computer interviews. InLREC 2014 May (pp. 3123–3128)

2. DeVault, D., Artstein, R., Benn, G., Dey, T., Fast, E., Gainer, A., Georgila, K., Gratch, J., Hartholt, A., Lhommet, M., Lucas, G., Marsella, S., Morbini, F., Nazarian, A., Scherer, S., Stratou, G., Suri, A., Traum, D., Wood, R., Xu, Y., Rizzo, A., and Morency, L.-P. (2014). “SimSensei kiosk: A virtual human interviewer for healthcare decision support”. In Proceedings of the 13th International Conference on Autonomous Agents and Multiagent Systems (AAMAS’14), Paris

3. Cuijpers P, Karyotaki E, Eckshtain D, Ng MY, Corteselli KA, Noma H, et al. Psychotherapy for depression across different age groups: A systematic review and meta-analysis. JAMA Psychiatry. 2020;77(7):694–702. doi:10.1001/jamapsychiatry.2020.0164

4. Daros AR, Haefner SA, Asadi S, Kazi S, Rodak T, Quilty LC. A meta-analysis of emotional regulation outcomes in psychological interventions for youth with depression and anxiety. Nat Hum Behav. 2021;5(10):1443–1457. doi:10.1038/s41562-021-01191-9

5. Dong B, Li B, Fan X, et al. A network analysis study of anxiety, depression and loneliness among middle-aged and elderly people in Xining area. BMC Psychol. 2025;13:931. doi:10.1186/s40359-025-03248-0

6. Fava GA. Can long-term treatment with antidepressant drugs worsen the course of depression? J Clin Psychiatry. 2003;64(2):123–133.

7. Fitzpatrick KK, Darcy A, Vierhile M. Delivering cognitive behavior therapy to young adults with symptoms of depression and anxiety using a fully automated conversational agent (Woebot): A randomized controlled trial. JMIR Ment Health. 2017;4(2):e19. doi:10.2196/mental.7785

8. Gururaj G, Varghese M, Benegal V, et al. *National Mental Health Survey of India*, *2015–16: Summary*. Bengaluru: National Institute of Mental Health and Neuro-Sciences (NIMHANS); 2016.

9. Kroenke K, Spitzer RL, Williams JB. The PHQ-9: Validity of a brief depression severity measure. J Gen Intern Med. 2001;16(9):606–613. doi:10.1046/j.1525-1497.2001.016009606.x

10. Kroenke K, Strine TW, Spitzer RL, Williams JB, Berry JT, Mokdad AH. The PHQ-8 as a measure of current depression in the general population. J Affect Disord. 2009;114(1–3):163–173. doi:10.1016/j.jad.2008.06.026

11. Lin, L.; Chen, X.; Shen, Y.; Zhang, L. Towards Automatic Depression Detection: A BiLSTM/1D CNN-Based Model. Appl. Sci. 2020, 10, 8701. 10.3390/app10238701

12. Lipton ZC. The mythos of model interpretability. Commun ACM. 2018;61(12):36–43. doi:10.1145/3233231

13. Megreya AM, Al-Emadi AA. The impact of cognitive emotion regulation strategies on math and science anxieties with or without controlling general anxiety. Sci Rep. 2024;14:19726. doi:10.1038/s41598-024-70705-y

14. Ministry of Health and Family Welfare, Government of India. National Telemedicine Service (eSanjeevani). New Delhi; 2020.

15. Nykoniuk M, Basystiuk O, Shakhovska N, Melnykova N. Multimodal data fusion for depression detection approach. Computation. 2025;13(1):9. doi:10.3390/computation13010009

16. Ohata W, Tani J. Investigation of the sense of agency in social cognition based on frameworks of predictive coding and active inference: A simulation study on multimodal imitative interaction. Front Neurorobot. 2020;14:61. doi:10.3389/fnbot.2020.00061

17. Patel V, Saxena S, Lund C, et al. The Lancet Commission on global mental health and sustainable development. Lancet. 2018;392(10157):1553–1598. doi:10.1016/S0140-6736(18)31612-X

18. Pavlov C, Egan K, Limbers C. Reliability and validity of the PHQ-8 in first-time mothers who used assisted reproductive technology. Hum Reprod Open. 2022;(2):hoac019. doi:10.1093/hropen/hoac019

19. Peng P, Chen Q, Liang M, Liu Y, Chen S, Wang Y, et al. A network analysis of anxiety and depression symptoms among Chinese nurses in the late stage of the COVID-19 pandemic. Front Public Health. 2022;10:996386. doi:10.3389/fpubh.2022.996386

20. Peters L, Peters A, Andreopoulos E, Pollock N, Pande RL, Mochari-Greenberger H. Comparison of DASS-21, PHQ-8, and GAD-7 in a virtual behavioral health care setting. Heliyon. 2021;7(3):e06473. doi:10.1016/j.heliyon.2021.e06473

21. Spitzer RL, Kroenke K, Williams JB. Validation and utility of a self-report version of PRIME-MD: The PHQ primary care study. JAMA. 1999;282(18):1737–1744. doi:10.1001/jama.282.18.1737

22. Tomitaka S, Kawasaki Y, Ide K, Akutagawa M, Yamada H, Furukawa TA. Item response patterns on the Patient Health Questionnaire-8 in a nationally representative sample of U.S. adults. Front Psychiatry. 2017;8:251. doi:10.3389/fpsyt.2017.00251

23. World Health Organization. Depression and Other Common Mental Disorders: Global Health Estimates. Geneva: WHO; 2017.

24. World Health Organization. Mental Health: Strengthening Our Response. Geneva: WHO; 2020.

25. Zeng Z, Pantic M, Roisman GI, Huang TS. A survey of affect recognition methods: Audio, visual, and spontaneous expressions. In: Proceedings of the 9th International Conference on Multimodal Interfaces; 2007. p. 126–133. doi:10.1145/1322192.1322212

